# Serum antibodies to surface proteins of *Chlamydia trachomatis* as candidate biomarkers of disease: Results from the Baltimore Chlamydia Adolescent/Young Adult Reproductive Management (CHARM) cohort

**DOI:** 10.1101/2021.05.25.21257614

**Authors:** Patricia X. Marques, Handan Wand, Melissa Nandy, Chun Tan, Huizhong Shou, Mishka Terplan, Katrina Mark, Rebecca M. Brotman, David P. Wilson, Jacques Ravel, Ru-ching Hsia, Patrik M. Bavoil

## Abstract

**Background:** We previously observed that the nine-member family of autotransported polymorphic membrane proteins (Pmps) of *Chlamydia trachomatis* is variably expressed in cell culture. Additionally, *C. trachomatis*-infected patients display variable Pmp-specific serum antibody profiles indirectly suggesting expression of unique Pmp profiles is an adaptive response to host-specific stimuli during infection. Here, we propose that the host response to Pmps and other outer surface proteins may correlate with disease severity.

**Methods:** This study tests this hypothesis using an ELISA that measures serum IgG antibodies specific for the nine *C. trachomatis* Pmp subtypes and four immunodominant antigens (MOMP, OmcB, GroEL, ClpP) in 265 participants of the *Chlamydia* Adolescent/Young Adult Reproductive Management (CHARM) cohort.

**Results:** More *C. trachomatis*-infected females displayed high Pmp-specific antibody levels (cut-off Indexes) than males (35.9-40.7% of females *vs*. 24.2-30.0% of males), with statistical significance for PmpC, F and H (P<0.05). Differences in Pmp-specific antibody profiles were not observed between *C. trachomatis*-infected females with a clinical diagnosis of pelvic inflammatory disease (PID) and those without. However, a statistically significant association between high levels of OmcB-specific antibody and a PID diagnosis (P<0.05) was observed.

**Conclusions:** Using antibody levels as an indirect measure of antigen expression, our results suggest that gender- and/or site-specific (cervix in females *vs*. urethra in males) stimuli control *pmp* expression in infected patients. They also support the possible existence of immune biomarkers of chlamydial infection associated with disease and underline the need for high resolution screening in human serum.

## INTRODUCTION

In 2018, more than 1.8 million cases of sexually transmitted *Chlamydia trachomatis* infection in the United States were reported to the Centers for Disease Control and Prevention (1). Chlamydial infections, which are often asymptomatic, can lead to pelvic inflammatory disease (PID), a precursor of female infertility, ectopic pregnancy, and chronic pelvic pain in 10-20% of infected women. Screening for *C. trachomatis* using nucleic acid amplification tests followed by treatment, can reduce the incidence of PID by as much as 60% (2), suggesting that detection of an active chlamydial infection is an effective means of reducing future sequelae. To fully control chlamydial disease however, host or chlamydial biomarkers that can discriminate between different clinical outcomes early in, or during, infection remain highly desirable.

A panoply of secreted and/or surface proteins of *C. trachomatis* are thought to act early in infection (3-6) as adhesins and/or invasins. These include the elementary body (EB)-specific cysteine-rich outer membrane complex protein OmcB (5), the major outer membrane protein MOMP (OmpA) (7), and members of the polymorphic membrane protein family (Pmps) (8). The late-expressed cysteine-rich protein OmcB was previously suggested as a candidate adhesin for *C. trachomatis* (9), *Chlamydia caviae* (3), and more recently for *Chlamydia pneumoniae* (5). MOMP displays sequence-variable domains defining 17 serovars categorized according to tissue tropism (A-C, ocular; D-K, urogenital; L1-L3, inguinal lymph nodes), and was also proposed to function as an adhesin for *C. trachomatis* (10). The 9-member *pmp* gene family of *C. trachomatis* (Pmp subtypes A-I; (11)) is predicted to be associated with tissue tropism (12). *C. trachomatis* PmpD-specific antibody has been shown to dramatically reduce infection *in vitro* (6, 13). These properties and the observed on/off switching of expression of the Pmps in cell culture (8, 14) suggest that production of a specific Pmp subtype(s) may be required and/or selected for at different sites or stages of infection. Consistent with variable expression in cell culture, we have previously shown that all *C. trachomatis*-infected patients mount a strong serologic response against different subsets of Pmps (15) with a suggested correlation between disease (PID) and PmpI antibody titer (15, 16).

The presence at the chlamydial surface of specific OmcB and MOMP types combined with the expression of diverse PmpA-I profiles may provide the outer coat diversity that is necessary for adherence to, internalization by and colonization of diverse mucosal sites. Since sites of infection may be associated with a spectrum of pathologies ranging from none to severe, we further propose that distinct outer surface protein profiles may also correlate with disease severity. Here we indirectly test these two linked hypotheses using an antibody capture ELISA to evaluate the host response to OmcB and MOMP, and as a surrogate for the expression of specific Pmp subtypes. We use serum samples from participants in the *Chlamydia* Adolescent/Young Adult Reproductive Management (CHARM) cohort enrolled in the Baltimore, Maryland area. To address site/gender specific expression, sera from *C. trachomatis*-infected men and women are compared. To address the possible link with disease severity, sera from women with a lower genital tract *C. trachomatis* infection and a reported PID diagnosis are compared with those without a PID diagnosis. Because of their previously reported association with disease in other studies, the *C. trachomatis* Heat Shock Protein GroEL (Hsp60) (17-19) and the Caseinolytic protease P (ClpP) (19) are also included.

## METHODS

### CHARM cohort

The <u>Ch</u>lamydia <u>A</u>dolescent & Young Adult <u>R</u>eproductive <u>M</u>anagement (CHARM) cohort consists of 276 *Chlamydia trachomatis*-infected men and women recruited between September 2010 and December 2013 in the Baltimore area (Table 1). CHARM inclusion and exclusion criteria, informed consent, treatment, data and specimens collected at entry and at intervals, follow-up activities, and statistical analyses were published independently (20). CHARM (HP-00042320) was reviewed and approved by the Institutional Review Board of the University of Maryland Baltimore. For the purpose of this study, blood was collected without additives at enrollment and prior to antibiotic treatment. After centrifugation, serum samples were stored at −20°C until analyzed. PID diagnosis was based on the minimum clinical criteria (cervical motion tenderness, uterine tenderness or adnexal tenderness) as set by the Centers for Disease Control and Prevention (21).

**TABLE 1:** Demographics and sexual behavior compared between genders

|  | Overall | Male N=120 (%) | Female N=145 (%) | P-value<sup>a</sup> |
| --- | --- | --- | --- | --- |
| Age, median (IQR) | 19.5 (17-21) | 20.2 (18-22) | 18.9 (17-21) | **0.0047<sup>b</sup>** |
| Age groups |  |  |  | **0.001** |
| ≤20 years old | 150 (56.60) | 55 (45.83) | 95 (65.52) |  |
| >20 years old | 115 (43.40) | 65 (54.17) | 50 (34.48) |  |
| **Sexual history** |  |  |  |  |
| Age of sexual debut |  |  |  | **0.007** |
| <13 years old | 223 (84.15) | 93 (77.50) | 130 (89.66) |  |
| 13+ years old | 42 (15.85) | 27 (22.50) | 15 (10.34) |  |
| Sexual partners lifetime |  |  |  | **<0.001** |
| <6 partners | 94 (35.47) | 26 (21.67) | 68 (46.90) |  |
| 6+ partners | 171 (64.53) | 94 (78.33) | 77 (53.10) |  |
| Oral sex | 233 (87.92) | 109 (90.83) | 124 (85.52) | 0.186 |
| Anal sex | 79 (29.81) | 41 (34.17) | 38 (26.21) | 0.159 |
| Condom use in past 3 months | 32 (11.99) | 21 (17.50) | 11 (7.59) | **0.014** |
| **Self-reported STI and reproductive health history** |  |  |  |  |
| Bacterial STI<sup>c</sup> | 206 (77.74) | 88 (73.33) | 118 (81.38) | 0.117 |
| Viral STI<sup>d</sup> | 27 (10.19) | 9 (7.50) | 18 (12.41) | 0.188 |
| *Trichomonas vaginalis* | N/A<sup>e</sup> | N/A | 28 (19.31) | N/A |
| BV | N/A | N/A | 15 (10.34) | N/A |
| PID | N/A | N/A | 6 (4.14) | N/A |
| **Physician assessment<sup>f</sup>** |  |  |  |  |
| PID | N/A | N/A | 33 (22.76) | N/A |
| **STI Diagnostic Laboratory tests (current)** |  |  |  |  |
| HIV | 18 (6.79) | 8 (6.67) | 10 (6.90) | 0.941 |
| **RPR/ syphilis** | 2 (0.75) | 1 (0.83) | 1 (0.69) | 0.893 |
| **GC/ gonorrhoea** | 20 (7.55) | 10 (8.33) | 10 (6.90) | 0.659 |

<sup>a</sup>P-values from chi-square, unless otherwise indicated
<sup>b</sup>P-values from Rank-sum Wilcoxon
<sup>c</sup>gonorrhoea, chlamydia and syphilis
<sup>d</sup>HPV, Herpes and HIV
N/A, not applicable, diagnosed in women only
PID assessment was performed by a physician following CDC criteria

### Recombinant polypeptides

The predicted passenger domains of PmpC, D, E, F, H and I, smaller fragments of PmpA (*pmpA-F2)*, PmpB (*pmpB-F1* and *pmpB–F2*) and PmpG (Figure 1) (15), rMOMP rGroEL, rOmcB and rClpP (from *C. trachomatis* serovar D; kindly provided by Dr. Guangming Zhong) were expressed in *E. coli* BL21(DE3) and purified as indicated in Table S1 (*Supplemental Digital Content 1 listing characteristics and purification conditions of the recombinant polypeptides*).

**FIGURE 1:**
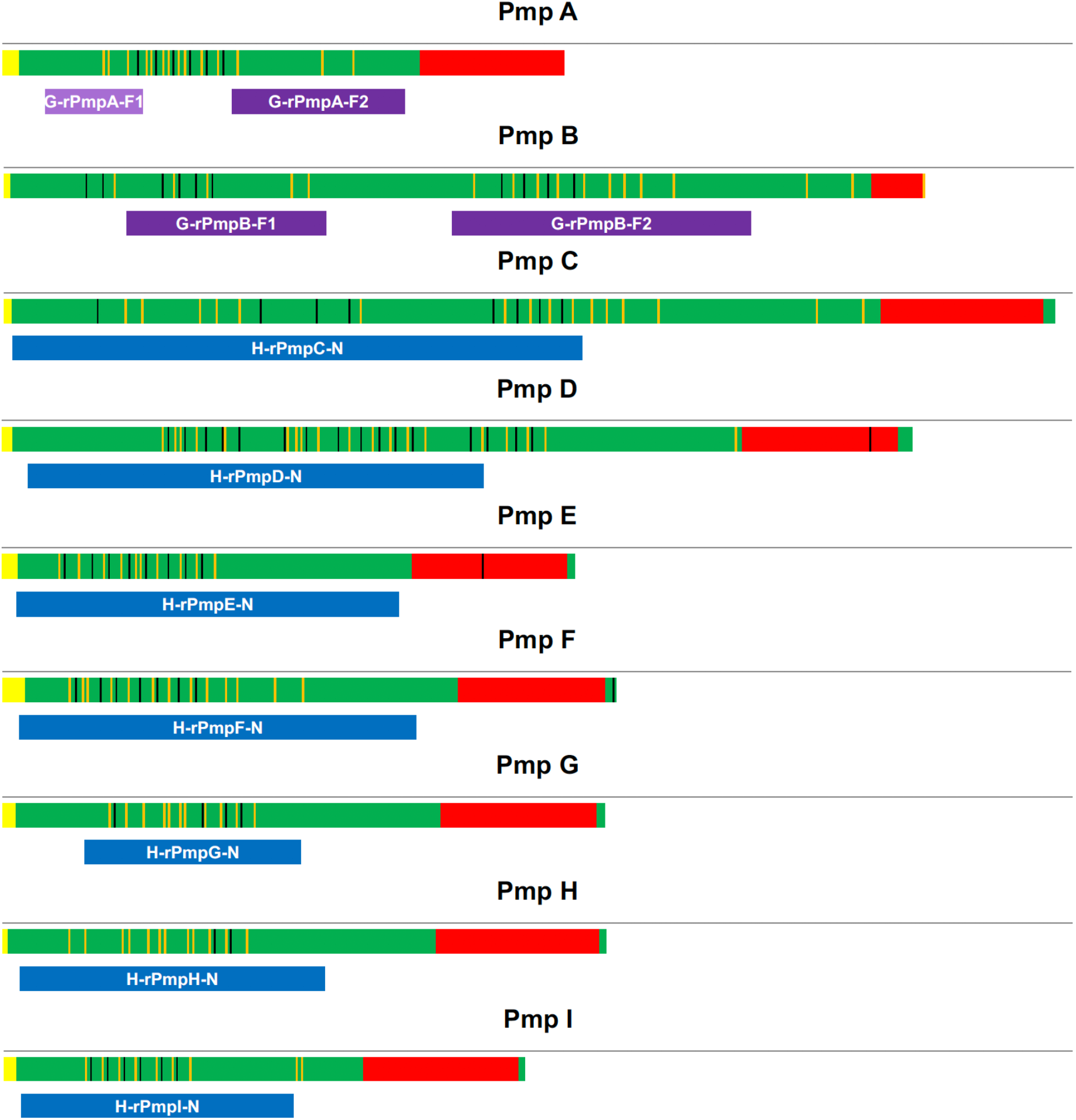
Schematic representation of the Pmp proteins and recombinant polypeptides used in this study. Structural features of Pmps are shown; Predicted signal peptide using Signal 4.0 software (yellow), passenger domain (green), GGA[I/L/V] (black bars) and FXXN (orange bars) tetrapeptide repeats and autotransporter domain (red). Recombinant Pmp derivatives based on predicted hydrophilic and antigenic domains located within the Pmp passenger domain, expressed in GST•tag pGEX-2t vector (GE Healthcare Biosciences) are shown in light purple (G-rPmpA-F1) and purple (G-rPmpA-F2, G-rPmpB-F1 and G-rPmp-F2). Passenger domain recombinant proteins expressed as inclusion bodies in His•tag pET30 (a or b) are in blue (H-rPmp(C to I)-N).

Insoluble polypeptides (all except rGroEL) were solubilized overnight at 42°C in 8 M urea/10 mg/ml Octyl beta-D-glucopyranoside (OGP) in buffer (50 mM Tris-HCl, pH 8/1 mM EDTA/1 mM DTT), dialyzed against 0.01% OGP and affinity purified (*Table S1, in Supplemental Digital Content 1)*.

Purification was monitored by SDS-PAGE and immunoblot (*see Figure S1, in Supplemental Digital Content 1, which shows Expression and purification of all the recombinant proteins and peptides used in this study)*.

## ELISA

Microplates PS (F-form; Greiner Bio-One North America, Inc.) coated with EBs purified from 48h *C. trachomatis* serovar E cultures and *C. pneumoniae* AR39 (22) (*see Table S2 in Supplemental Digital Content 1, that lists the ELISA optimized conditions*) or purified recombinant polypeptides *(see Table S2, in Supplemental Digital Content 1*) were incubated overnight at 4°C, washed with PBS-T and blocked with 2.5% milk. ELISA was performed as described (23) using diluted CHARM sera (*Table S2, Supplemental Digital Content 1*) and goat anti-human IgG peroxidase labeled antibody (1:2000 dilution; 100 µl/well; KPL, Inc.). Results were read in a Beckman DTX 880 plate reader (Beckman Coulter, Inc.) at 450 and 620 nm (background).

Results obtained by ELISA were comparable to those previously obtained by immunoblot analysis and densitometry (15) (not shown), thus supporting the validity of results obtained by either method. ELISA results generated with GST•tagged and His•tagged OmcB were positively correlated (p<0.001; *see Figure S2, in Supplemental Digital Content 1, a graphic of the correlation between G-OmcB and H-OmcB*), further validating the ELISA.

### Statistical analysis

Chi-squared test was used to compare the demographic, sexual behavior and biological factors of the study groups. Median and inter-quartile ranges were used to describe continuous variables. A non-parametric test was used to compare the median by gender.

Antibody levels defined as the cut-off Indexes for rPmpA to I, rClpP, rHsp60, rMOMP, rOmcB and *C. trachomatis* purified EBs were used as categorical variables and split into tertiles. The first tertile cut-point represented the lower, while the third tertile represented the higher cut-off indexes. Logistic regression models were then used to determine the association between cut-off indexes in the first and third tertiles (highest/lowest) and PID diagnosis (outcome). Unadjusted and adjusted Odds Ratios (ORs) and their 95% confidence intervals (CIs) were calculated. In adjusted analysis, results were presented after accounting for potential confounding effects of age, *Trichomonas vaginalis* infection and a diagnosis of bacterial vaginosis (BV) based on Amsel’s criteria (24).

All analyses were performed using Stata 12.0 (College Station).

## RESULTS

### Demographics and sexual behavior in the CHARM cohort

A total of 265 participants (120 males, 45.3% and 145 females, 54.7%) of the CHARM cohort were included in the study. The overall median age of the study population was 19.5 years old (IQR: 17-21). Males were more likely to be older (median: 20.2 years, IQR: 18-22) compared to women (18.9 years, IQR: 17-21 respectively) (p=0.0047). Table 1 compares demographic data, sexual behavior, sexual history and past and current diagnosis of sexually transmitted infections (STIs) in men and women of the CHARM cohort (20). Most of the study population reported having first vaginal intercourse before age 13 years (84.2%). Females were more likely to report a sexual debut before age 13 years (86.7 females vs. 77.5%, males, p=0.007). The majority of the participants (64.5%) reported having 6 or more lifetime sexual partners. Male participants were more likely than females to report 6+ sexual partners (53.1% females vs. 78.3% males, p<0.001). There were no significant differences reported for receptive oral or anal sex based on gender. Of the 145 CHARM female participants whose samples were analyzed by ELISA, 126 (86.9%) were heterosexual, one (0.7%) was homosexual and 18 (12.4%) were bisexual. Thirty seven (25.5%) reported receptive anal sex. Of the CHARM 120 male participants, 105 (87.5%) were heterosexual, 9 (7.5%) were homosexual and 6 (5%) bisexual. Thirteen (10.8%) reported receptive anal sex. Overall, 12.0% of the participants reported condom use in the last 3 months and females were significantly less likely to report having sex with partners who used condoms compared to males (7.6% females *vs*. 17.5% males, p=0.014). Most of the CHARM participants were African Americans or of Hispanic/Latino ethnicity.

### Factors associated with pelvic inflammatory disease (PID) in the females of the CHARM cohort

Of the 145 CT+ female participants from the CHARM cohort, 33 (22.8%) were clinically diagnosed as having PID at the study visit (Table 2). We assessed associations between a PID diagnosis with various demographic and sexual risk factors. Participants self-reporting previous *T. vaginalis* infection (OR: 2.79, 95% CI: 1.15 – 6.80, p-value=0.0230.025), and previous PID (OR: 19.82, 95% CI: 2.23 – 176.52, p-value=0.007) were more likely to be diagnosed with PID at study entry.

**TABLE 2:**
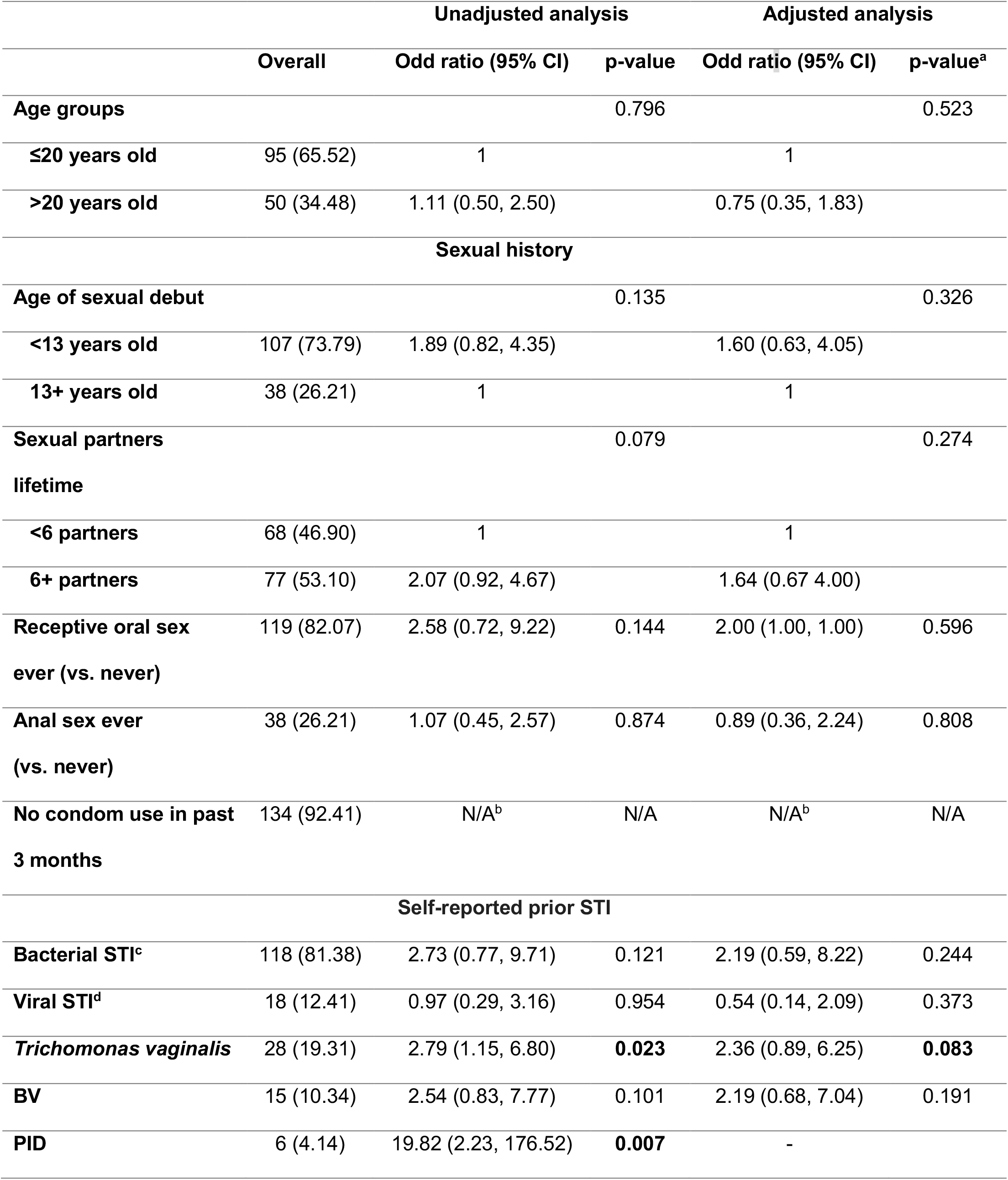
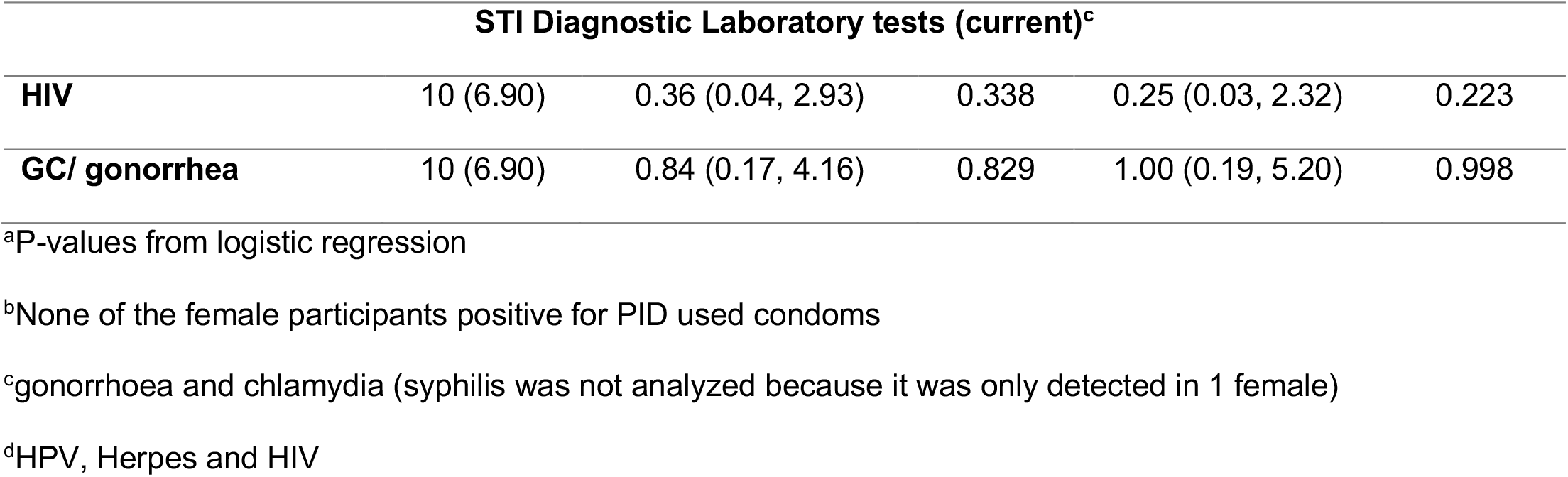
Association of demographic, biological and sexual behaviors in 33 women with a PID diagnosis among the 145 women in the study

### The serum Pmp-specific antibody profile of *C. trachomatis*-infected CHARM participants is associated with gender

*C. trachomatis* and *C. pneumoniae* purified EBs and recombinant antigen ELISA were used to analyze serum antibodies (IgG) of 265 patients (145153 females and 120 males) from the CHARM cohort. Most participants had an antibody response against H-rOmcB, H-rPmpC, G-rHsp60, and H-rPmpI with sensitivity above 85% (*Table S3, Supplemental Digital Content 1*). The antibody response against *C. pneumoniae* purified EBs was very low with only 3 samples (1.1%) with slightly elevated antibody levels.

Potential association between serum antibody levels (grouped in tertiles) and gender was investigated. Overall, when compared to males, a higher proportion of females were in the top tertile across all Pmps (35.9-40.7% for females vs. 24.2-30.0% for males; Table 3). Significant gender differences were observed for rPmps subtypes H-rPmpC-N (39.3% vs. 25.8%, p=0.001 for females and males respectively), H-rPmpF-N (36.6% vs 29.2%, p=0.020) and rPmpH-N (40.7% vs 24.2%, p=0.018). Trending statistical significance was observed for G-rPmpA-F2 (39.3% vs. 25.8%, p=0.068), and H-rPmpE-N (39.3% vs. 25.8%, p=0.063). For G-MOMP, a significant proportion of males were in the lower tertile when compared to females (26.2% vs. 42.5%, p=0.011). Gender-based differences were not detected for other recombinant peptides.

**TABLE 3:** *Chlamydia* antigenic biomarkers compared between genders (categorized by tertiles)

|  | Overall | Male N=122 (%) | Female N=145 (%) | P-value<sup>a</sup> |
| --- | --- | --- | --- | --- |
| **G-rPmpA-F2** |  |  |  | **0.068** |
| <0.41 | 89 (33.58) | 45 (37.50) | 44 (30.34) |  |
| 0.41 – 1.04 | 88 (33.21) | 44 (36.67) | 44 (30.34) |  |
| >1.04 | 88 (33.21) | 31 (25.83) | 57 (39.31) |  |
| **G-rPmpB-F1** |  |  |  | 0.586 |
| <0.28 | 89 (33.58) | 43 (35.83) | 46 (31.72) |  |
| 0.28 – 0.69 | 88 (33.21) | 41 (34.17) | 47 (32.41) |  |
| >0.69 | 88 (33.21) | 36 (30.00) | 52 (35.86) |  |
| **G-rPmpB-F2** |  |  |  | 0.417 |
| <0.55 | 90 (33.96) | 42 (35.00) | 48 (33.10) |  |
| 0.55 – 2.40 | 87 (32.83) | 43 (35.83) | 44 (30.34) |  |
| >2.40 | 88 (33.21) | 35 (29.17) | 53 (36.55) |  |
| **H-rPmpC-N** |  |  |  | **0.001** |
| <8.75 | 89 (33.58) | 54 (45.00) | 35 (24.14) |  |
| 8.75 – 20.47 | 88 (33.21) | 35 (29.17) | 53 (36.55) |  |
| >20.47 | 88 (33.21) | 31 (25.83) | 57 (39.31) |  |
| **H-rPmpD-N** |  |  |  | 0.195 |
| <1.88 | 89 (33.58) | 47 (39.17) | 42 (28.97) |  |
| 1.88 – 4.10 | 88 (33.21) | 38 (31.67) | 50 (34.48) |  |
| >4.10 | 88 (33.21) | 35 (29.17) | 53 (36.55) |  |
| **H-rPmpE-N** |  |  |  | **0.063** |
| <6.13 | 89 (33.58) | 46 (38.33) | 43 (29.66) |  |
| 6.13 – 14.94 | 88 (33.21) | 43 (35.83) | 45 (31.03) |  |
| >14.94 | 88 (33.21) | 31 (25.83) | 57 (39.31) |  |
| **H-rPmpF-N** |  |  |  | **0.020** |
| **<5.52** | 89 (33.58) | 51 (42.50) | 38 (26.21) |  |
| **5.52 – 19.85** | 88 (33.21) | 34 (28.33) | 54 (37.24) |  |
| **>19.85** | 88 (33.21) | 35 (29.17) | 53 (36.55) |  |
| **H-rPmpG-N** |  |  |  | **0.199** |
| **<0.79** | 89 (33.58) | 44 (36.67) | 45 (31.03) |  |
| **0.79 – 3.69** | 88 (33.21) | 43 (35.83) | 45 (31.03) |  |
| **>3.69** | 88 (33.21) | 33 (27.50) | 55 (37.93) |  |
| **H-rPmpH-N** |  |  |  | **0.018** |
| **<1.93** | 89 (33.58) | 46 (38.33) | 43 (29.66) |  |
| **1.93 – 5.17** | 88 (33.21) | 45 (37.50) | 43 (29.66) |  |
| **>5.17** | 88 (33.21) | 29 (24.17) | 59 (40.69) |  |
| **H-rPmpI-N** |  |  |  | **0.105** |
| **<4.19** | 89 (33.58) | 46 (38.33) | 43 (29.66) |  |
| **4.19 – 10.68** | 88 (33.21) | 42 (35.00) | 46 (31.72) |  |
| **>10.68** | 88 (33.21) | 32 (26.67) | 56 (38.62) |  |
| **G-rClpP** |  |  |  | **0.709** |
| **<0.68** | 89 (33.58) | 39 (32.50) | 50 (34.48) |  |
| **0.68 – 2.08** | 88 (33.21) | 38 (31.67) | 50 (34.48) |  |
| **>2.08** | 88 (33.21) | 43 (35.83) | 45 (31.03) |  |
| **G-rGroEL** |  |  |  | **0.952** |
| **<4.11** | 89 (33.58) | 40 (33.33) | 49 (33.79) |  |
| **4.11 – 13.58** | 88 (33.21) | 39 (32.50) | 49 (33.79) |  |
| **>13.58** | 88 (33.21) | 41 (34.17) | 47 (32.41) |  |
| **G-rMOMP** |  |  |  | **0.011** |
| **<1.64** | 89 (33.58) | 51 (42.50) | 38 (26.21) |  |
| **1.64 – 4.61** | 88 (33.21) | 32 (25.83) | 57 (39.31) |  |
| **>4.61** | 88 (33.21) | 38 (31.67) | 50 (34.48) |  |
| **G-rOmcB** |  |  |  | 0.830 |
| **<1.94** | 89 (33.58) | 40 (33.33) | 49 (33.79) |  |
| **1.94 – 4.44** | 88 (33.21) | 42 (35.00) | 46 (31.72) |  |
| **>4.44** | 88 (33.21) | 38 (31.67) | 50 (34.48) |  |
| **H-rOmcB** |  |  |  | 0.835 |
| **<7.00** | 89 (33.58) | 38 (31.67) | 51 (35.17) |  |
| **7.00 – 24.83** | 88 (33.21) | 41 (34.17) | 47 (32.41) |  |
| **>24.83** | 88 (33.21) | 41 (34.17) | 47 (32.41) |  |
| **Ct purified EBs<sup>b</sup>** |  |  |  | 0.308 |
| **<1.96** | 89 (33.58) | 43 (35.83) | 46 (31.72) |  |
| **1.96 – 5.22** | 88 (33.21) | 43 (35.83) | 45 (31.03) |  |
| **>5.22** | 88 (33.21) | 34 (28.33) | 54 (37.24) |  |

<sup>a</sup>P-values from chi-square
<sup>b</sup>*Chlamydia trachomatis* serovar E purified EBs

**TABLE 4:** Assessing associations between the levels of the factors (in tertiles) associated with PID diagnosis (tertiles calculated only for females, described on Table S4)

| Biomarkers | second vs first |  | third vs first |  |
| --- | --- | --- | --- | --- |
|  | Unadjusted Odds Ratio (95% CI) | p-value | Unadjusted Odds Ratio (95% CI) | p-value<sup>a</sup> |
| G-rPmpA-F2 | 0.66 (0.26 – 1.67) | 0.379 | 0.58 (0.22 – 1.50) | 0.258 |
| G-rPmpB-F1 | 0.64 (0.24 – 1.67) | 0.362 | 0.82 (0.33 – 2.08) | 0.680 |
| G-rPmpB-F2 | 1.83 (0.70 – 4.75) | 0.214 | 1.17 (0.43 – 3.19) | 0.760 |
| H-rPmpC-N | 0.92 (0.36 – 2.34) | 0.855 | 0.81 (0.31 – 2.11) | 0.667 |
| H-rPmpD-N | 1.03 (0.41- 2.59) | 0.954 | 0.71 (0.27 – 1.89) | 0.494 |
| H-rPmpE-N | 1.77 (0.70 – 4.47) | 0.225 | 0.78 (0.28 – 2.18) | 0.636 |
| H-rPmpF-N | 0.71 (0.27 – 1.89) | 0.494 | 1.03 (0.41 – 2.59) | 0.954 |
| H-rPmpG-N | 1.57 (0.63 – 3.89) | 0.330 | 0.59 (0.21 – 1.68) | 0.322 |
| H-rPmpH-N | 0.66 (0.26 – 1.67) | 0.379 | 0.58 (0.22 – 1.50) | 0.258 |
| H-rPmpl-N | 2.33 (0.88 – 6.16) | **0.088** | 1.35 (0.48 – 3.77) | 0.569 |
| G-ClpP | 0.92 (0.37 – 2.29) | 0.863 | 0.55 (0.21 – 1.49) | 0.242 |
| G-rGroEL | 1.48 (0.56 – 3.93) | 0.429 | 1.48 (0.56 – 3.93) | 0.429 |
| G-rMOMP | 1.48 (0.56 – 3.93) | 0.429 | 1.48 (0.56 – 3.93) | 0.429 |
| G-rOmcB | 2.95 (1.03 – 8.49) | **0.045** | 2.66 (0.92 – 7.72) | **0.072** |
| H-rOmcB | 2.62 (0.83 – 8.21) | **0.099** | 4.83 (1.61 – 14.47) | **0.005** |
| Ct purified EBs<sup>b</sup> | 1.52 (0.55 – 4.20) | 0.415 | 2.11 (0.79 – 5.63) | 0.135 |
|  | second vs first |  | third vs first |  |
|  | Adjusted<sup>c</sup> Odds Ratio (95% CI) | p-value | Adjusted<sup>c</sup> Odds Ratio (95% CI) | p-value |
| G-rPmpA-F2 | 0.68 (0.26 – 1.79) | 0.432 | 0.76 (0.27 – 2.14) | 0.606 |
| G-rPmpB-F1 | 0.80 (0.28 – 2.25) | 0.668 | 0.98 (0.35 – 2.69) | 0.962 |
| G-rPmpB-F2 | 2.00 (0.72 – 5.52) | 0.181 | 1.01 (0.35 – 2.92) | 0.983 |
| H-rPmpC-N | 0.98 (0.36 – 2.66) | 0.974 | 0.85 (0.31 – 2.33) | 0.752 |
| **H-rPmpD-N** | 1.07 (0.41 – 2.81) | 0.892 | 0.86 (0.31 – 2.40) | 0.774 |
| **H-rPmpE-N** | 2.21 (0.80 – 6.10) | 0.128 | 1.06 (0.35 – 3.19) | 0.922 |
| **H-rPmpF-N** | 0.77 (0.28 – 2.16) | 0.622 | 1.38 (0.51 – 3.75) | 0.529 |
| **H-rPmpG-N** | 1.96 (0.74 – 5.17) | 0.175 | 0.68 (0.22 – 2.03) | 0.486 |
| **H-rPmpH-N** | 0.74 (0.28 – 1.98) | 0.551 | 0.71 (0.25 – 1.99) | 0.510 |
| **H-rPmpI-N** | 2.52 (0.89 – 7.11) | **0.081** | 1.77 (0.59 – 5.36) | 0.310 |
| **G-ClpP** | 0.80 (0.31 – 2.11) | 0.657 | 0.51 (0.18 – 1.45) | 0.208 |
| **G-rGroEL** | 1.28 (0.47 – 3.49) | 0.634 | 1.40 (0.51 – 3.87) | 0.517 |
| **G-rMOMP** | 1.32 (0.48 – 3.63) | 0.592 | 1.39 (0.50 – 3.85) | 0.529 |
| **G-rOmcB** | 2.58 (0.87 – 7.63) | **0.087** | 2.56 (0.86 – 7.65) | **0.091** |
| **H-rOmcB** | 2.31 (0.72 – 7.46) | 0.160 | 4.22 (1.37 – 13.01) | **0.012** |
| **Ct purified EBs<sup>c</sup>** | 1.59 (0.56 – 4.57) | 0.385 | 2.23 (0.80 – 6.24) | 0.127 |

<sup>a</sup> P-values from logistic regression
<sup>b</sup> *Chlamydia trachomatis* serovar E purified EBs
<sup>c</sup> Adjusted for age, sexual partners ever, *Trichomonas vaginalis* and Bacterial Vaginosis (BV)

### The serum OmcB-specific antibody response is associated with a clinical diagnosis of pelvic inflammatory disease (PID) in *C. trachomatis*-infected female CHARM participants

High antibody levels against H-rOmcB (Figure 2; Table *4 and see S4, in Supplemental Digital Content 1, showing associations between factors (in tertiles) associated with PID diagnosis with tertiles calculated only for females*; OR: 2.31, 95% CI: 0.72-7.46, p=0.160 and OR: 4.22, 95% CI: 1.37-13.01, p=0.012, respectively) were significantly associated with a clinical diagnosis of PID. Similar results were obtained for G-rOmcB (Figure 2; Table*s 4 and S4) associated with PID diagnosis with tertiles calculated only for females*; OR: 2.58, 95% CI: 0.87-7.63, p=0.087/second vs. first tertile, OR: 2.56, 95% CI: 0.86-7.65, p=0.091/third vs. first tertile).

**FIGURE 2:**
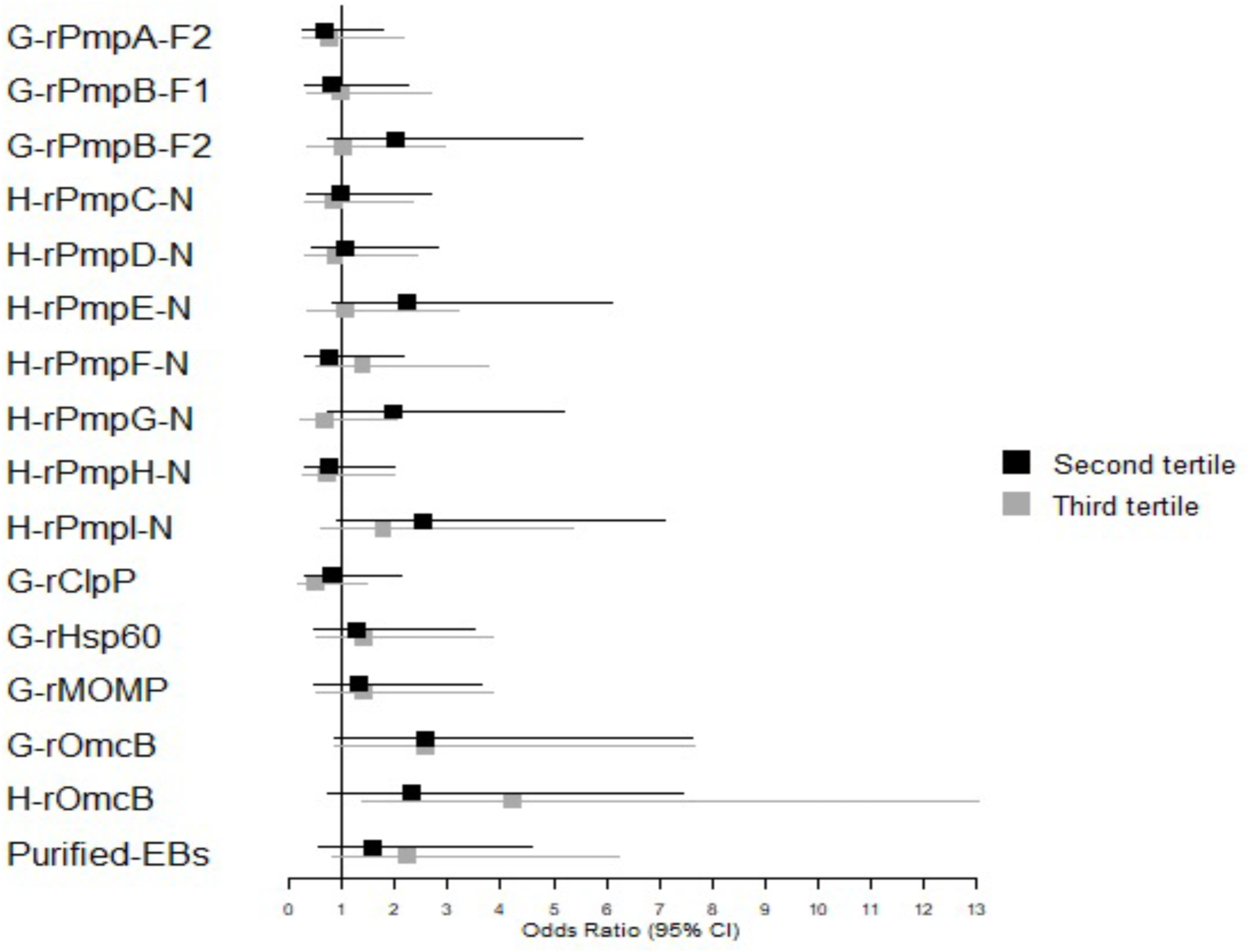
High cut-off index antibodies associated with a PID diagnosis. Sera from 153 female CHARM participants were analyzed by ELISA. Odds ratios (ORs) for PID positivity when a patient is in the second (black) or third (grey) tertile, compared to the first tertile are shown. The OR (95% confidence interval) was greater than 1 for G-rOmcB in both tertiles and H-rOmcB in the second tertile (* p<0.05) suggesting that high antibody cut-off indexes for this antigen were significantly associated with PID. Results for H-rPmpI-N second tertile were borderline significant (‡ p=0.075).

In contrast, antibodies to the other tested antigens were not individually associated with PID with any statistical significance. Antibody levels for the second and third tertiles were almost identical for Hsp60 and MOMP (Figure 2.). Similarly, antibody levels against each of the Pmp subtypes were not significantly associated with PID, although the response to H-rPmpI was borderline significant (Figure 2; OR: 2.52, 95% CI: 0.89-7.11, p=0.075/first vs. second tertile). Interestingly, however, antibody levels against 8 of the 9 rPmp subtypes (all but H-rPmpF) were slightly higher in women without a clinical diagnosis of PID, although none reached statistical significance individually. This suggests a global negative association of antibodies against the Pmp family and upper genital tract disease.

## DISCUSSION AND CONCLUSIONS

An ELISA was developed to measure antibody levels against recombinant polypeptides derived from the 9 *C. trachomatis* Pmps, the EB-specific OmcB, MOMP, ClpP, and the stress response protein GroEL, in 265 chlamydia-positive male and female participants of the CHARM cohort from the Baltimore area. All participants exhibited an antibody response to *C. trachomatis* purified EBs and at least one of the Pmps. While female CHARM participants had proportionally higher antibody levels to all Pmps compared to males, statistically significant gender differences were observed for 3 Pmps (rPmpC, F and H). The higher antibody levels in females are consistent with previous reports of sex differences in antibody responses (reviewed in (25)). However, since *C. trachomatis* infection in men and women is diagnosed at different sites (the cervix in women and urethra in men), these findings are also consistent with the hypothesis that different Pmps may be expressed differentially in different anatomical sites and/or physiological environments.

Statistically significant differences in anti-Pmp antibody levels were not detected when comparing female CHARM participants with or without a positive clinical diagnosis for PID, indicating that the Pmp-specific antibody response is not significantly altered when the infection ascends to the upper genital tract compared to when it is limited to the lower genital tract. In turn, this suggests that the expressed Pmp profile of *C. trachomatis-* infected women is independent of the site of infection along the reproductive tract. Although Pmp-based differences were not observed, CHARM participants with a clinical PID diagnosis were more likely to have high antibody levels against OmcB than those without. This is consistent with previous reports of OmcB being associated with CD8+ T cell-mediated upper genital tract immunopathology (26). In contrast, antibody levels were almost identical for GroEL and MOMP, suggesting that these antigens are not associated with upper genital tract infection and PID, consistent with previous findings for GroEL (27) and MOMP (28). Our study failed to confirm the reported association of ClpP-specific antibodies with upper genital tract disease (19). However, this earlier study involved a different end point (tubal factor infertility) and a smaller cohort.

Overall, the observation of different antibody responses to specific Pmp subtypes in *C. trachomatis*-infected patients relative to gender and/or site of infection, while compatible with the hypothesized site-specific expression of Pmps in the context of infection, falls short of providing indisputable evidence for it, and does not lend confidence that these antigens are exploitable for sero-diagnosis. Multiple behavioral, immunological and structural factors have historically confounded or limited serologic analyses of chlamydial antigens for diagnostic purposes. For instance, potential cross-reactivity with orthologous antigens owing to *Chlamydia pneumoniae* (29) or *Chlamydia psittaci* (30) previous or concurrent infections, gender-associated differential immune responses (25), co-infection with other STIs, repeat infection with *C. trachomatis* or concurrent infection at multiple sites, as well as non-native structural features of recombinant antigens, are potential limitations of any approach toward reliable sero-diagnosis. Conversely, any statistically significant serologic difference that is detectable above the background ‘noise’ generated by these confounding factors and limitations should be exploitable as a diagnostic or prognostic tool. In this context, our findings that OmcB-specific serum antibody levels were elevated in women with a PID diagnosis suggests that this antigen is worthy of further study as a biomarker of upper genital tract disease. For instance, the possibility of determining whether an asymptomatic *C. trachomatis*-infected woman is likely to develop or already has subclinical upper genital tract pathology (silent PID; (31)) via a simple measurement of OmcB-specific antibody would be of great benefit to the physician and ultimately to his/her patient. Although cross-reactive antibody responses against *C. pneumoniae* OmcB have been reported (32), Gijsen et al. (33) also showed that tubal pathology was more common in patients with both *C. pneumoniae* and *C. trachomatis* antibodies compared to patients with antibodies against OmcB from only one *Chlamydia* species, further strengthening a role for this well-conserved chlamydial protein in eliciting pathology.

In conclusion, our expanded analysis of the observed Pmp-specific antibody profile variation in *C. trachomatis*-infected men and women of the CHARM cohort may reflect the hypothesized differential expression of different Pmp profiles as a prerequisite or in response to infection of distinct urogenital sites. However other possibilities exist that suggest that these antigens may be impractical to use as serologic biomarkers. Our study also revealed an association between a high OmcB-specific serum antibody level and a PID diagnosis. This finding corroborates previous studies (18) and indicates that this protein may provide an exploitable discriminatory biomarker of disease severity during *C. trachomatis* infection.

## Data Availability

All data provided in the manuscript are available upon request.

## Acknowledgements

The contributions of Esther Colinetti, Nyaradzo Longinaker, Anup Mahurkar and Dr. Ligia Peralta to the development of the CHARM cohort are gratefully acknowledged.

## Supplemental Digital Content 1.pptx

**Supplemental Digital Content 1. Table S1, which lists the** characteristics and purification conditions of the recombinant polypeptides.

**Supplemental Digital Content 1. Figure S1, showing Expression and purification of all the recombinant proteins and peptides used in this study. A)** Coomassie staining of all recombinant proteins; 1: G-rPmpA-F2, 2: G-rPmpB-F1, 3: G-rPmpB-F2, 4 to 10: H-rPmp(C to I)-N, 11: G-rClpP, 12: G-rHsp60, 13: G-rMOMP, 14: G-rOmcB, 15: H-rOmcB and 16: GST only. **B)** Immunoblots. Primary antibodies: guinea pig anti-Pmp A to I (lanes 1-10), mouse anti-ClpP (provided by Dr. Guangming Zhong) (lane 11), rabbit anti-GroEL (lane 12) and anti-OmcB (lanes 14-15) and goat anti-MOMP (LifeSpan BioSciences) (lane 13) and anti-GST (GE Healthcare Bio-Sciences) (lane 16); secondary antibodies: Alkaline phosphatase (AP)-conjugated anti-guinea pig, -mouse (KPL, Inc.), -rabbit and –goat (Sigma-Aldrich). Membranes were revealed with fluorescent alkaline phosphatase substrate ECF (GE Healthcare Biosciences). *Apparent molecular weights of protein bands are listed in Table S1.

**Supplemental Digital Content 1. Table S2**, listing ELISA optimized conditions

**Supplemental Digital Content 1. Figure S2, Correlation between G-OmcB and H-OmcB**. OmcB recombinant protein was obtained from 2 different clones using GST•tag pGEX-2t vector (G-OmcB) and His•tag pET30a vector (H-OmcB). Positive correlation between the ELISA results obtained with these 2 proteins was statistical significant (0.7462, p<0.001).

**Supplemental Digital Content 1. Table S3**, showing the ELISA results, sensitivity and coefficient variation

**Supplemental Digital Content 1. Table S4** listing tertiles for females only (n=153)

## Notes

Research reported in this publication was supported by the National Institute of Allergy and Infectious Diseases of the National Institutes of Health under award number U19AI084044. The content is solely the responsibility of the authors and does not necessarily represent the official views of the National Institutes of Health

### Competing Interest Statement

The authors have declared no competing interest.

### Funding Statement

Research reported in this publication was supported by the National Institute of Allergy and Infectious Diseases of the National Institutes of Health under award number U19AI084044. The content is solely the responsibility of the authors and does not necessarily represent the official views of the National Institutes of Health. No other services or payment were received from a third party by any of the authors or their institutions

### Author Declarations

CHARM (HP-00042320) was reviewed and approved by the Institutional Review Board of the University of Maryland Baltimore. The CHARM study details are described in reference 20: Mark KS, Brotman RM, Martinez-Greiwe S, Terplan M, Bavoil P, Ravel J. Chlamydia in adolescent/adult reproductive management trial study (CHARM): Clinical core protocol. Contemp Clin Trials Commun. 2019;16:100414

