## Supplemental files for "Serum antibodies to surface proteins of *Chlamydia trachomatis* as candidate biomarkers of disease: Results from the Baltimore Chlamydia Adolescent/Young Adult Reproductive Management (CHARM) cohort"

Table S1: Marques *et al.*

**Table S1:** Characteristics and purification conditions of the recombinant polypeptides.

| Recombinant fragment | Vector | Amino acid residues | Molecular mass (kDa) Calc[a] | App[b] | Equilibration Buffer | Detergent and urea | Purification column |
|---|---|---|---|---|---|---|---|
| **G-rPmpA-F2** | pGEX-2t | 314-604 | 58[c] | 45 | PBS, pH 7.2 | Triton X-100, OGP and urea | Glutathione (GST-Tag) |
| **G-rPmpB-F1** | pGEX-2t | 208-542 | 61.3[c] | 70 | PBS, pH 7.2 | Triton X-100, OGP and urea | Glutathione (GST-Tag) |
| **G-rPmpB-F2** | pGEX-2t | 754-1256 | 77.8[c] | 75 | PBS, pH 7.2 | Triton X-100, OGP and urea | Glutathione (GST-Tag) |
| **H-rPmpC-N** | pET30a | 16-974 | 105 | 100 | Sodium phosphate, pH 7 | Triton X-100, OGP and urea | Talon (His-Tag) |
| **H-rPmpD-N** | pET30a | 45-810 | 78.6 | 75 | Sodium phosphate, pH 7 | Triton X-100, OGP and urea | Talon (His-Tag) |
| **H-rPmpE-N** | pET30a | 26-667 | 80.6 | 80 | Sodium phosphate, pH 7 | Triton X-100, OGP and urea | Talon (His-Tag) |
| **H-rPmpF-N** | pET30b | 29-697 | 79.3 | 75 | PBS, pH 7.2 | Triton X-100, OGP and urea | Talon (His-Tag) |
| **H-rPmpG-N** | pET30a | 139-502 | 43.1 | 45 | Sodium phosphate, pH 7 | Triton X-100, OGP and urea | Talon (His-Tag) |
| **H-rPmpH-N** | pET30c | 30-542 | 63.6 | 65 | Sodium phosphate, pH 7 | Triton X-100, OGP and urea | Talon (His-Tag) |
| **H-rPmpI-N** | pET30b | 30-488 | 55.4 | 55 | PBS, pH 7.2 | Triton X-100, OGP and urea | Talon (His-Tag) |
| **G-rClpP** | pGEX-6p-2 | 1-192 | 47.2[a] | 50 | PBS, pH 7.2 | Triton X-100. OGP and urea | Glutathione (GST-Tag) |
| **G-rGroEL** | pGEX-2t | 1-544 | 84.1[a] | 85 | PBS, pH 7.2 | Triton X-100 | Glutathione (GST-Tag) |
| **G-rMOMP** | pGEX-2t | 32-280 | 53.4[a] | 54 | PBS, pH 7.2 | Triton X-100, OGP and urea | Glutathione (GST-Tag) |
| **G-rOmcB** | pGEX-2t | 92-553 | 75.8[a] | 75 | Sodium phosphate, pH 7 | Triton X-100, OGP and urea | Glutathione (GST-Tag) |
| **H-rOmcB** | pET30a | 92-553 | 49.6 | 50 | Sodium phosphate, pH 7 | Triton X-100, OGP and urea | Talon (His-Tag) |

[a]Calc: Molecular mass (kDa) calculated using ProtoParam tool (ExPaSy – Bioinformatics resource portal)

[b] App: Molecular mass (kDa) apparent based on Figure S2

[c] +26 kDa (GST-tag)

Figure S2: Marques *et al.*

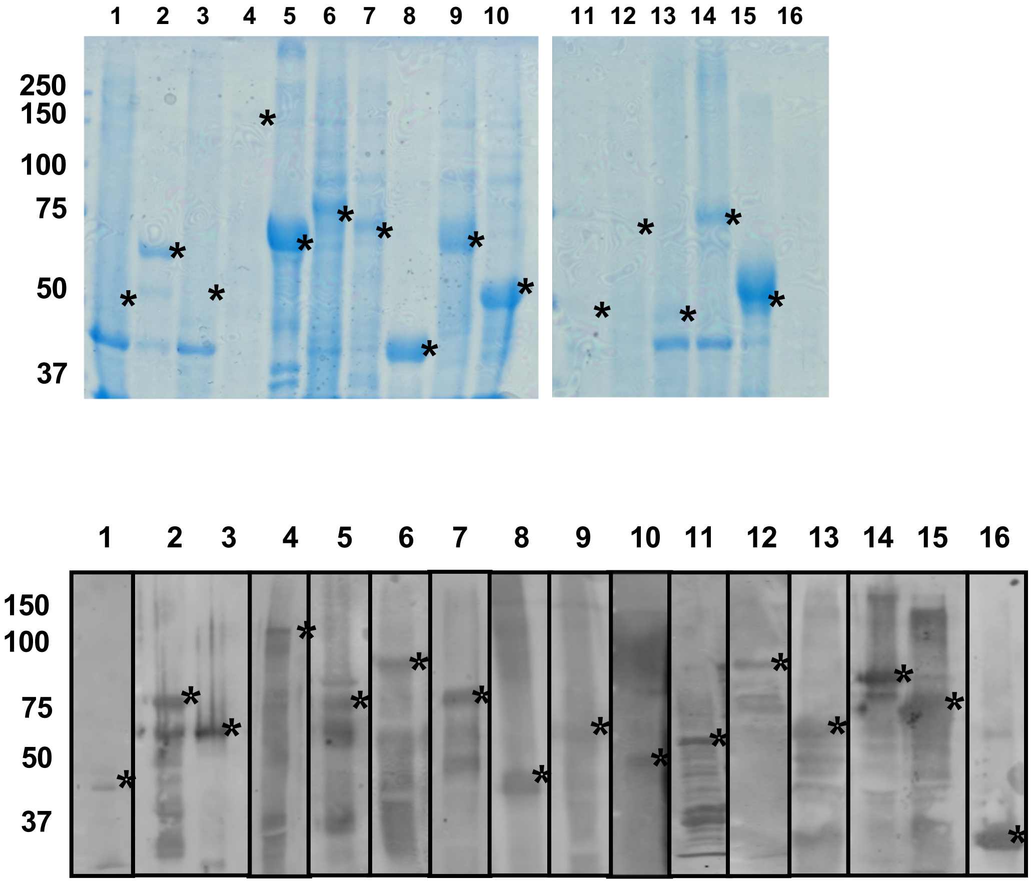

Table S2: Marques *et al.*

**Table S2:** ELISA optimized conditions

| Recombinant peptide and EBs | Cut-off[a] | Concentration of purified recombinant peptides used for ELISA coating (µg/ml) | Serum dilution[b] |
|---|---|---|---|
| **G-rPmpA-F2** | 0.240 | 20 | 1:200 |
| **G-rPmpB-F1** | 0.306 | 20 | 1:200 |
| **G-rPmpB-F2** | 0.056 | 40 | 1:200 |
| **H-rPmpC-N** | 0.031 | 10 | 1:100 |
| **H-rPmpD-N** | 0.039 | 20 | 1:100 |
| **H-rPmpE-N** | 0.0204 | 40 | 1:100 |
| **H-rPmpF-N** | 0.011 | 20 | 1:100 |
| **H-rPmpG-N** | 0.073 | 40 | 1:100 |
| **H-rPmpH-N** | 0.121 | 40 | 1:100 |
| **H-rPmpI-N** | 0.032 | 10 | 1:100 |
| **G-rClpP** | 0.146 | 10 | 1:100 |
| **G-rGroEL** | 0.04 | 10 | 1:200 |
| **G-rMOMP** | 0.117 | 10 | 1:200 |
| **G-rOmcB** | 0.164 | 10 | 1:200 |
| **H-rOmcB** | 0.136 | 5 | 1:800 |
| **Ct purified EBs[c]** | 0.072 | 5 | 1:50 |
| **Cpn purified EBs[d]** | 0.072 | 5 | 1:50 |
| **rGST** | N/A[e] | 5 | 1:200 |

[a]Cut-offs calculated using 27 Ct negative serum samples (Mean +3*Standard Deviation)
[b]Different dilutions were used in some samples
[c]*Chlamydia trachomatis* serovar E purified EBs
[d]*Chlamydia pneumoniae* AR39 purified EBs
[e]N/A: not applicable

Figure S3: Marques *et al.*

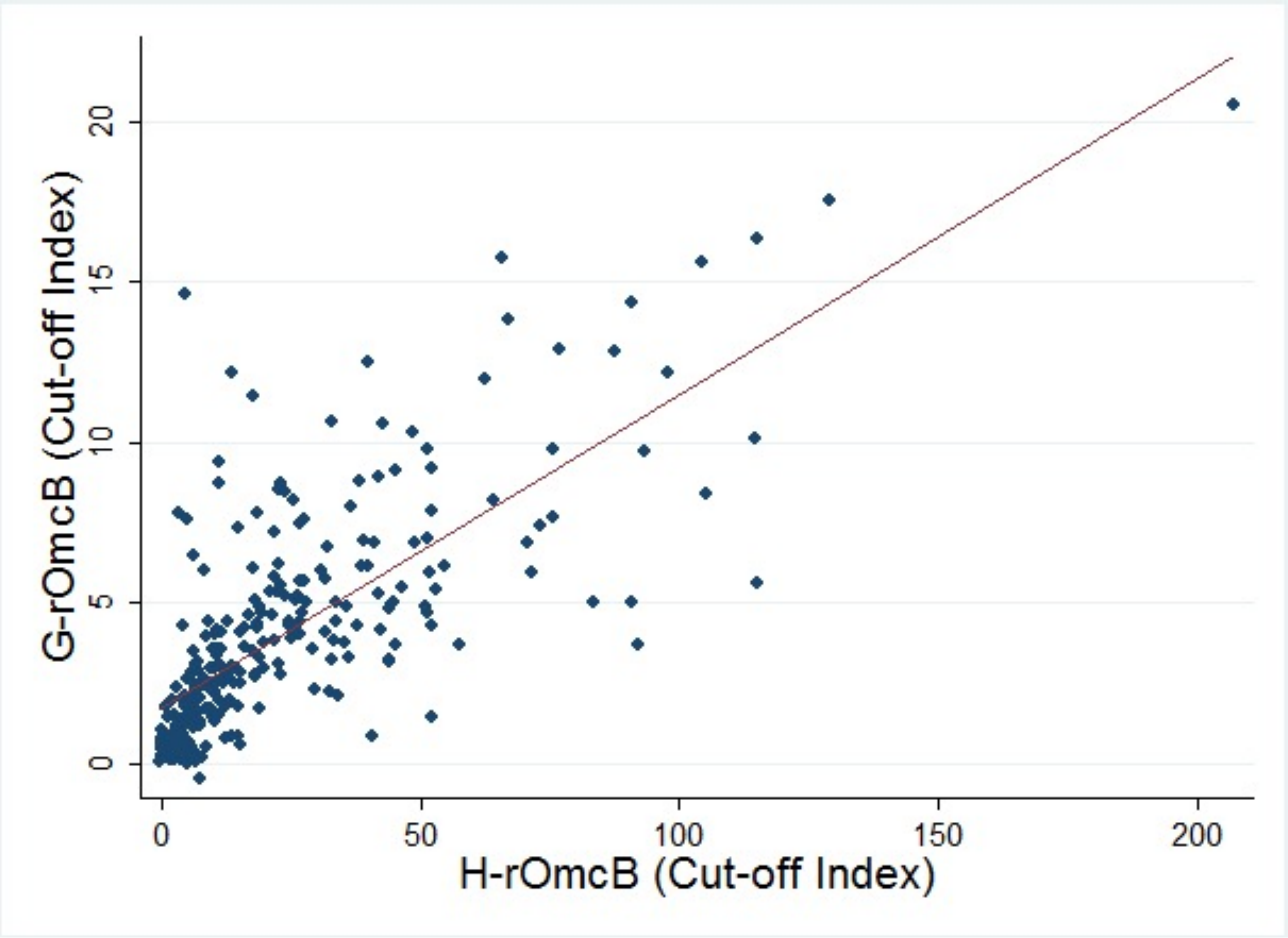

Table S3: Marques *et al.*

**Table S3:** ELISA results, sensitivity and coefficient variation

| | **Total (n=265)** | | **Female (n=145)** | | **Male (n=120)** | | **Sensitivity (%)** |
|---|---|---|---|---|---|---|---|
| | **Positive (%)** | **Negative (%)** | **Positive (%)** | **Negative (%)** | **Positive (%)** | **Negative (%)** | |
| **G-rPmpA-F2** | 83 (31.32) | 182 (68.68) | 55 (37.93) | 90 (62.07) | 28 (23.33) | 92 (76.67) | 60.26 |
| **G-rPmpB-F1** | 50 (18.87) | 215 (81.13) | 33 (22.76) | 112 (77.24) | 17 (14.17) | 103 (85.83) | 56.21 |
| **G-rPmpB-F2** | 130 (49.06) | 135 (50.94) | 74 (51.03) | 71 (48.97) | 56 (46.67) | 64 (53.33) | 67.15 |
| **H-rPmpC-N** | 248 (93.58) | 17 (6.42) | 140 (96.55) | 5 (3.45) | 108 (90.00) | 12 (10.00) | 94.20 |
| **H-rPmpD-N** | 204 (76.98) | 61 (23.02) | 114 (78.62) | 31 (21.38) | 90 (75.00) | 30 (25.00) | 81.90 |
| **H-rPmpE-N** | 200 (75.47) | 65 (24.53) | 100 (68.97) | 45 (31.03) | 100 (83.33) | 20 (16.67) | 80.94 |
| **H-rPmpF-N** | 202 (76.23) | 63 (23.77) | 118 (81.38) | 27 (18.62) | 84 (70.00) | 36 (30.00) | 81.42 |
| **H-rPmpG-N** | 167 (63.02) | 98 (36.98) | 97 (66.90) | 48 (33.10) | 70 (58.33) | 50 (41.67) | 73.80 |
| **H-rPmpH-N** | 187 (70.57) | 78 (29.43) | 109 (75.17) | 36 (24.83) | 78 (65.00) | 42 (35.00) | 77.97 |
| **H-rPmpI-N** | 223 (84.15) | 42 (15.85) | 125 (86.21) | 20 (13.79) | 98 (81.67) | 22 (18.33) | 86.79 |
| **G-rClpP** | 132 (49.81) | 133 (50.19) | 71 (48.97) | 74 (51.03) | 61 (50.83) | 59 (49.17) | 67.48 |
| **G-rGroEL** | 236 (89.06) | 29 (10.94) | 128 (88.28) | 17 (11.72) | 108 (90.00) | 12 (10.00) | 90.49 |
| **G-rMOMP** | 194 (73.21) | 71 (26.79) | 113 (77.93) | 32 (22.07) | 81 (67.50) | 39 (32.50) | 79.54 |
| **G-rOmcB** | 211 (79.62) | 54 (20.38) | 118 (81.38) | 27 (18.62) | 93 (77.50) | 27 (22.50) | 83.64 |
| **H-rOmcB** | 249 (93.96) | 16 (6.04) | 141 (97.24) | 4 (2.76) | 108 (90.00) | 12 (10.00) | 94.52 |
| **Ct purified EBs[a]** | 210 (79.25) | 55 (20.75) | 119 (82.07) | 26 (17.93) | 91 (75.83) | 29 (24.17) | 83.38 |
| **Cpn purified EBs[b]** | 3 (1.13) | 262 (98.87) | 2 (1.38) | 143 (98.62) | 1 (0.83) | 119 (99.17) | N/A[c] |

[a]*Chlamydia trachomatis* serovar E purified EBs
[b]*Chlamydia pneumoniae* AR39 purified EBs
[c]N/A: not applicable

**Table S5:** Tertiles for females only (n=153)

| | first tertile | second tertile | third tertile |
|---|---|---|---|
| **G-rPmpA-F2** | < 0.46 | 0.46 – 1.29 | > 1.29 |
| **G-rPmpB-F1** | < 0.29 | 0.29 – 0.74 | > 0.74 |
| **G-rPmpB-F2** | < 0.55 | 0.55 – 2.56 | > 2.56 |
| **H-rPmpC-N** | < 12.25 | 12.25 – 24.15 | > 24.15 |
| **H-rPmpD-N** | < 2.13 | 2.13 – 4.44 | > 4.44 |
| **H-rPmpE-N** | < 7.49 | 7.49 – 17.64 | > 17.64 |
| **H-rPmpF-N** | < 8.45 | 8.45 – 21.17 | > 21.17 |
| **H-rPmpG-N** | < 1.14 | 1.14 – 4.22 | > 4.22 |
| **H-rPmpH-N** | < 2.32 | 2.32 – 5.93 | > 5.93 |
| **H-rPmpI-N** | < 4.7 | 4.7 – 12.20 | > 12.20 |
| **G-rClpP** | < 0.66 | 0.66 – 2.04 | > 2.04 |
| **G-rGroEL** | < 3.74 | 3.74 – 13.53 | > 13.53 |
| **G-rMOMP** | < 2.04 | 2.04 – 4.72 | > 4.72 |
| **G-rOmcB** | < 1.93 | 1.93 – 4.67 | > 4.67 |
| **H-rOmcB** | < 6.66 | 6.66 – 24.61 | > 24.61 |
| **Ct purified EBs[a]** | < 2.07 | 2.07 – 6.31 | > 6.31 |

[a]*Chlamydia trachomatis* serovar E purified EBs
